# Reimagining the serocatalytic model for infectious diseases: a case study of common coronaviruses

**DOI:** 10.1101/2024.12.10.24318816

**Authors:** Sophie L. Larsen, Junke Yang, Huibin Lv, Yang Wei Huan, Qiwen Teo, Tossapol Pholcharee, Ruipeng Lei, Akshita B Gopal, Evan K. Shao, Logan Talmage, Chris K. P. Mok, Saki Takahashi, Alicia N. M. Kraay, Nicholas C. Wu, Pamela P. Martinez

**Author notes:** These authors contributed equally (Junke Yang, Huibin Lv).

## Abstract

Despite the increased availability of serological data, understanding serodynamics remains challenging. Serocatalytic models, which describe the rate of seroconversion (gain of antibodies) and seroreversion (loss of antibodies) within a population, have traditionally been fit to cross-sectional serological data to capture long-term transmission dynamics. However, a key limitation is their binary assumption on serological status, ignoring heterogeneity in optical density levels, antibody titers, and/or exposure history. Here, we implemented Gaussian mixture models - an established statistical tool - to cross-sectional data in order to characterize serological diversity of seasonal human coronaviruses (sHCoVs) throughout the lifespan. These methods identified four (NL63, 229E, OC43) to five (HKU1) distinct seropositive levels, suggesting that among seropositive individuals, the number of prior exposures or response to infection may vary. For each sHCoV, we fit adapted, multi-compartment serocatalytic models across 10 scenarios with different assumptions on exposure history and waning of antibodies. The best fit model for each sHCoV was always one that accounted for a gradient of seropositivity as well as host variation in the scale of serological response to infection. These models allowed us to estimate the strength and frequency of serological responses across sHCoVs, finding that the time for a seronegative individual to become seropositive ranges from 2.33-4.07 years across sHCoVs, and most individuals mount a strong antibody response reflected in high optical density values, skipping lower levels of seropositivity. We also find that despite frequent infection and strong serological responses, it is rare for an individual to remain seropositive throughout the lifetime. Crucially, our reimagined serocatalytic methods can be flexibly adapted across pathogens, having the potential to be broadly applied beyond this work.

## Introduction

Serological data, combined with robust statistical and mathematical methods, are widely used to infer key parameters guiding the behavior of pathogens [1]. When working with cross-sectional serological data, serocatalytic models are traditionally used, which divide the population into seronegative (no detected serological response to a pathogen) and seropositive (a serological response is present), and capture movement between these compartments across continuous age [1, 2]. This tool has long been used to address questions about infectious disease transmission (e.g. [3–7]). To account for heterogeneity in behavior or exposure risk, serocatalytic implementations often include stratification of the force of infection by age. However, age is only one of many sources of host heterogeneity in infectious disease (e.g.[8–11]), and one of the main limitations of serocatalytic models is that they collapse serological diversity to two compartments with binary seropositivity, ignoring the potential role of serological heterogeneity on disease transmission and control.

Seasonal human coronaviruses (sHCoVs) are an ideal pathogen system with which to test adapted serocatalytic approaches that incorporate heterogeneity in host exposure history. There are four known sHCoVs in circulation and they account for 15%-30% of common colds [12]. This widespread circulation in the human population [13, 14] not only motivates further investigation into their dynamics but also enables robust serological sampling across the lifespan that can inform modeling approaches. Additionally, their close relationship with SARS-CoV-2 has been leveraged throughout the pandemic to gain insight into potential future dynamics (e.g. [15–18]). Therefore, understanding sHCoV transmission and serodynamics can have broad implications for public health.

Here, we implemented Gaussian mixture models to characterize diversity in serology for the four sHCoVs, using cross-sectional samples from China covering 0-67 years of age [19]. After identifying patterns in this serological data, we expanded classical serocatalytic models to capture a range of hypotheses for the development of exposure history and serological variation. By fitting these models to data, we estimated seroconversion rates across a gradient of serostatus levels, including the scalar change in protection from reinfection for each class, as well as variation in the strength of serological responses to exposure. We also estimated the rate of maternal antibody loss and the median age of first infection for each sHCoV, which can inform the optimal timing of infant SARS-CoV-2 vaccination. The flexibility of this approach has the potential to be used for other pathogens of interest.

## Results

### Capturing serostatus heterogeneity

To capture the seroprevalence trajectories of sHCoVs across the lifespan, we analyzed the reactivity of plasma samples from individuals between age 0−67 (*n* = 2, 414) to the major antigens of the four sHCoVs, namely the spike S1 subunits of NL63, 229E, and HKU1, as well as the hemagglutinin esterase protein of OC43. Plasma reactivity was measured by ELISA based on the values of optical density at 450 nm (OD_450_).

The plasma reactivity data for donors between age 0−18 were obtained in our previous study [19], whereas all the data for adults (age *>* 18) were measured in the present study. To identify the threshold OD_450_ values between seronegative and seropositive samples in our data for all individuals, we applied 2-component Gaussian mixture models [20], which are commonly used for this purpose [1, 2], to data for infants under age one. This population subset was chosen to ensure capturing true seronegatives: maternal antibodies are likely to wane during this time period [21], and many have not yet had a natural infection [6]. With this approach, the OD_450_-value threshold for each sHCoV ranged from 0.408−0.513, and the range of seropositive OD_450_ values was wide (Figures 1A, S2). The widest range of OD_450_ values for seropositives was observed for HKU1-S1, varying from 0.422 to 2.651, representing a 6-fold change in OD_450_ values between the upper and lower values.

**Figure 1:**
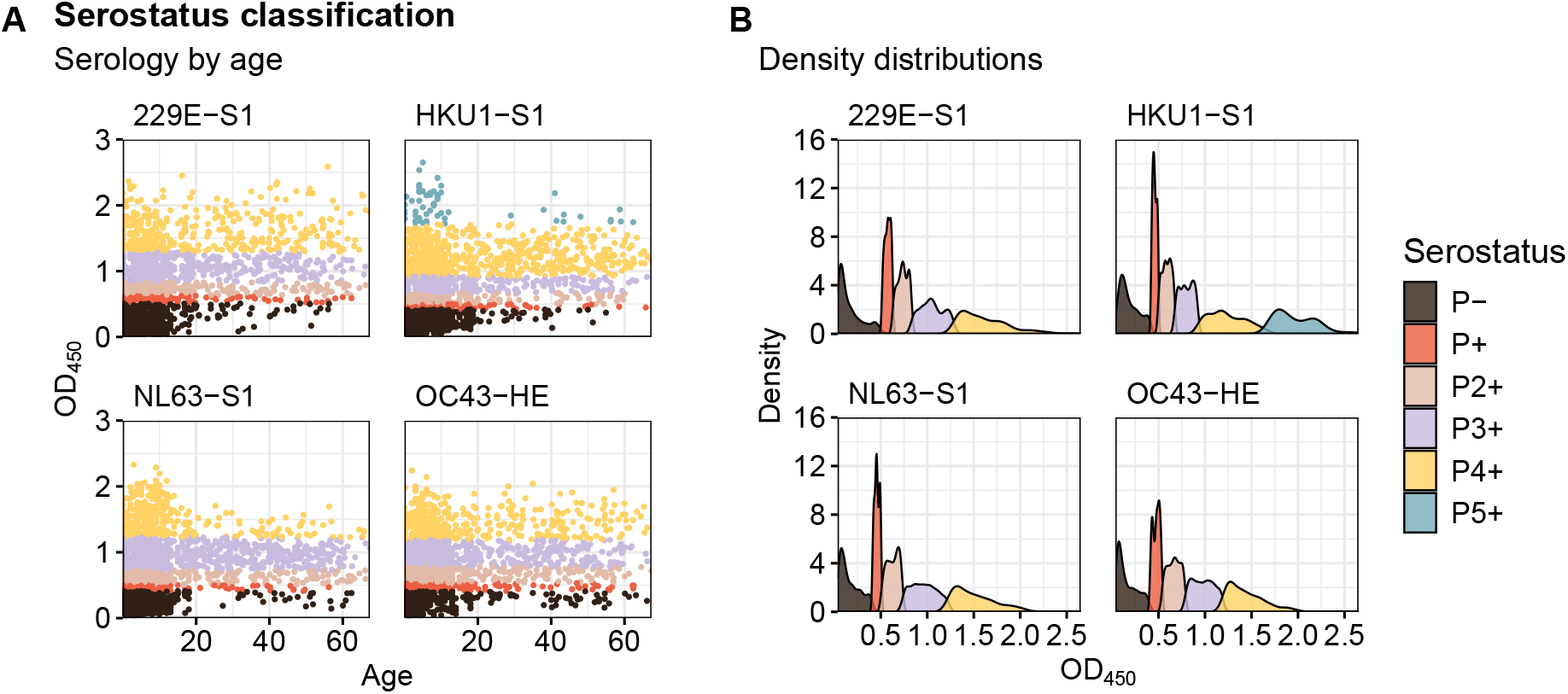
Distribution of OD_450_ values. (A) OD_450_ values by age under gradient classification. (B) OD_450_ probability density under gradient classification. Colors indicate the serostatus classification of 2,414 individuals based on the Gaussian mixture model. Four seropositive groups are identified for sHCoVs 229E, OC43, and NL63, and five for sHCoV HKU1 (Figure S1). Outcomes under the binary classification, as well as the frequency of OD_450_ values are shown in Figure S2.

To explore serological diversity and test whether seropositive samples could be further distinguished, we reran the mixture model on only seropositive samples, this time allowing the number of seropositive components to be inferred based on the Bayesian Information Criterion (Figures 1, S1). This approach returned mixture models with five seropositive components for HKU1-S1, and four for 229E-S1, NL63-S1, and OC43-HE. We refer to this as a gradient of seropositivity. When looking at OD_450_ values throughout the lifespan, the distribution of OD_450_ values at each serostatus level widens with increasing mean OD_450_ value (Figure 1), representing increased heterogeneity as serostatus level increases. In order to disentangle varying hypotheses for the development of gradient sHCoV seropositivity throughout the lifespan, we implemented mathematical models of seroconversion (via exposure) and seroreversion (loss of antibodies).

### Reimagining serocatalytic models

We first implemented a binary serocatalytic model with a compartment for maternal seropositives (individuals who are seropositive due to maternal antibodies rather than previous pathogen exposure) (Figure 2A, Equation S1). This model has traditionally been used to measure seroconversion rates from cross-sectional data that has been classified into seronegative and seropositive [1, 3], and it is referred here as the “Binary Model”. In this work, we consider seroconversion to occur from exposure only (no vaccination), and the time to seroconversion (1/rate) is measured as the time until getting infected plus the time to become detectably seropositive from an infection. In order to test potential mechanisms for the development of gradient seropositivity, we also developed more complex serocatalytic models that include a number of seropositive compartments corresponding to the gradient identified by the mixture models. In the first such gradient model, which we call the “Ordered Model”, seroconversion moves an individual to the next-highest serostatus level - for example, moving from *P*^−^ to *P*_+_ (Figure 2B-C, Equation S2). In the second of these gradient models, which we call the “Variation Model”, individuals can increase their serostatus level by one or more seropositive compartments - for example, *P*^−^ to *P*_4+_ (Figure 2D-E, Equation S3). For these two gradient models, we assumed that seroreversion either proceeded one serostatus level at a time (“Sequential Waning”, Figures 2B, D) or direct to seronegative (“Direct Waning”, Figures 2C, E) at a rate *ρ*. Compartments for maternal seropositivity are also included in the gradient models, for individuals whose maternal antibodies have not yet waned. For all models, candidate values for the seroreversion rate *ρ* were estimated from longitudinal data [22].

**Figure 2:**
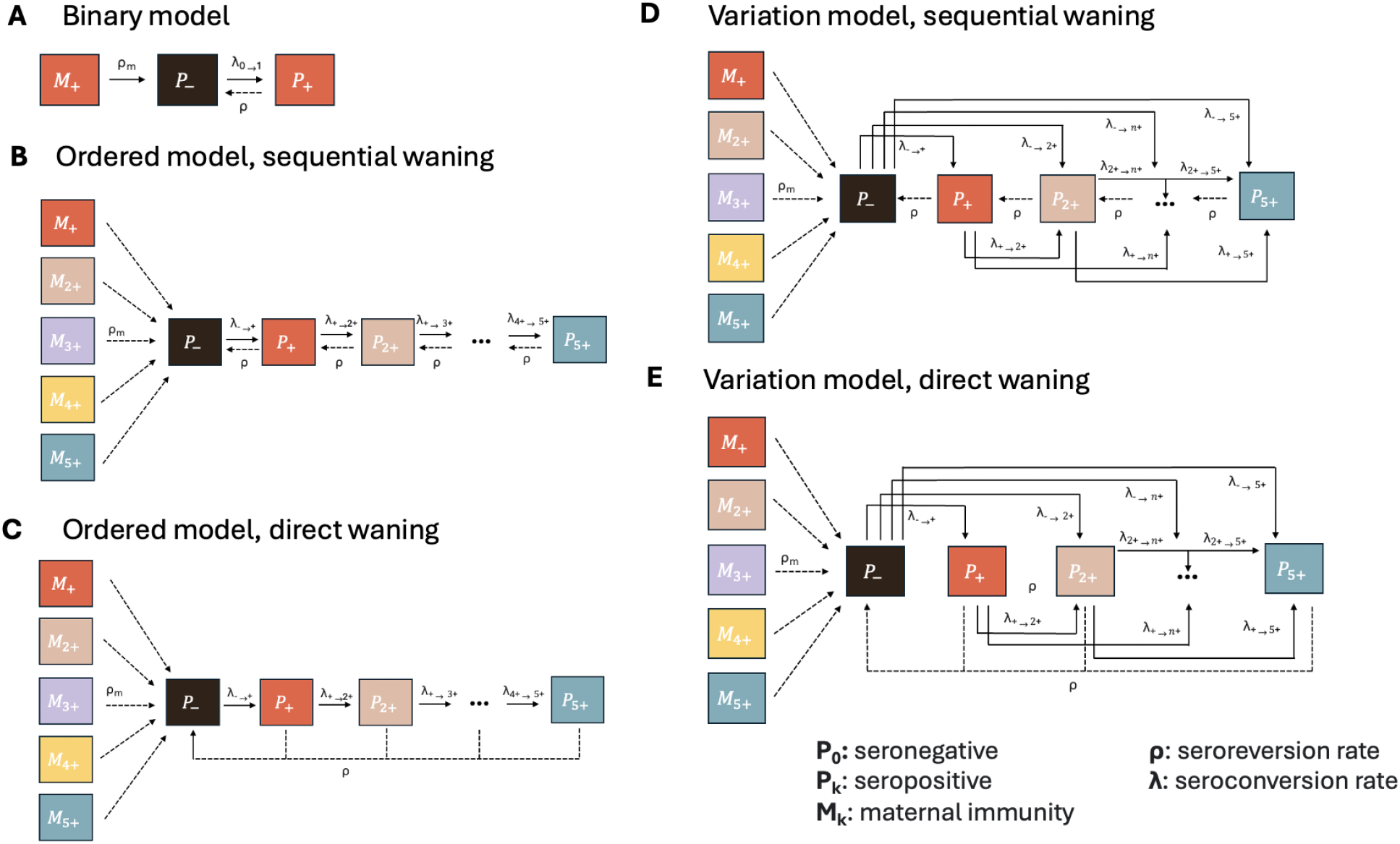
Model diagrams for all five model structures considered. (A) In the Binary Model, only seronegative (*P*_−_), seropositive (*P*_+_), and maternally seropositive (*M*_+_) compartments are considered. (B-C) In the Ordered Model, we incorporate a gradient of seropositive compartments (*M*_+_,*M*_2+_,…, *P*_+_, *P*_2_+, …) and individuals can seroconvert one serostatus level at a time, with higher levels indicating higher OD_450_ values. Seroreversion is “sequential” (B), moving individuals to successively lower serosatus levels, or “direct” (C), moving individuals directly to seronegative (*P*^−^). (D-E) The Variation Model includes the same gradient of seropositivity as the Ordered Model, but also includes heterogeneity in serological response to infection. In particular, individuals can seroconvert not just from *P*_*i*_ ⟶ *P*_*i*+1_ but also to *P*_*n*_ where *n* is any higher serostatus level. Sequential (D) and direct (E) seroreversion are considered. Individuals seroconvert at a rate *λ* and serorevert at a rate *ρ*.

When comparing these models with the Akaike Information Criterion (AIC), the Binary Model was always outperformed by models capturing gradient serostatus levels, regardless of the assumptions used for fitting (Figure 3). Across all four sHCoVs, we found that the best-performing set of assumptions was the Variation Model with a slow, sequential seroreversion rate (*ρ* = 0.39, Figure 3C). We also compared the models using the Root Mean Square Error (RMSE), finding a 2-to 3-fold difference in RMSE values favoring the Variation Model over the Binary Model (Figures S4). Finally, we profiled all fit parameters for the Binary Model and Variation Model, and there was no loss of identifiability in the Variation Model (Figure S6, Tables S4, S7).

**Figure 3:**
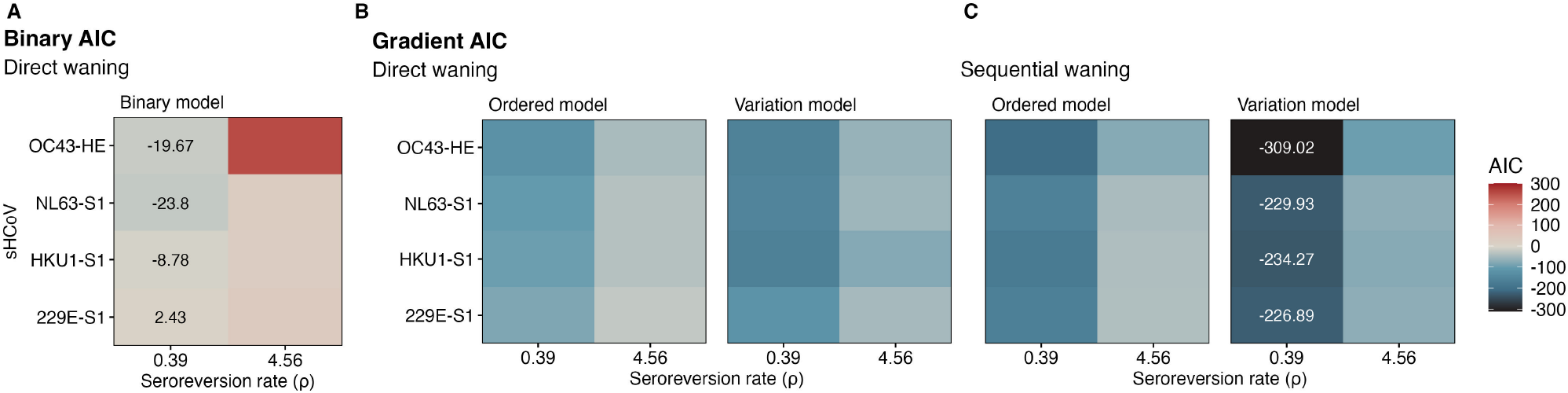
Identification of best-fit serocatalytic models. (A) Akaike Information Criterion (AIC) values across binary serostatus classifications. (B-C) AIC values across Ordered and Variation models are shown with direct (B) or sequential (C) waning. Two values for the seroreversion rate *ρ* are tested, based on estimates from longitudinal data [22], and the effects of these values on waning serostatus classes over time are compared in Figure S3. Lower AIC values indicate better model performance, and the AIC values of the best (white text) and worst (black text) performing models are shown for each sHCoV. Fit parameters are listed in Tables S1-S2, starting parameters and boundary values are shown in Table S4, and initial conditions are shown in Tables S5-S6.

### Disentangling exposure history

Having identified the best-performing model, we next sought to explore the implications for seroconversion and seroreversion rates and, more broadly, the development of exposure history. When looking at parameter estimates (Table 1), the maternal seroreversion rate (*ρ*_*m*_) was similar across sHCoVs with a range of 5.13 − 7.65 years^−1^, corresponding to 48 − 71 days of maternal seropositivity. When looking at seroconversion, we first estimated that seroconversion rates were similarly frequent across sHCoVs. For example, the estimated time to seroconversion for seronegative individuals (*λ*_−⟶*n*+_) ranged from 2.33 to 4.07 years. When comparing seroconversion rates across sHCoVs, all showed similar rates of seroconversion out of each serostatus level until *P*_3+_, where HKU1-S1, 229E-S1, and OC43-HE all showed frequent seroconversion (3.51 − 6.89 years) but NL63-S1 seroconversion was infrequent (one in a lifetime). These rates are also reflected in the trajectories of Figure 4A. For example, NL63-S1 serostatus trajectories stabilized in adulthood with *P*_3+_ as the dominant serostatus level. In contrast, *P*_3+_ and *P*_4+_ serostatus levels exhibited similar prevalence for 229E-S1 and OC43-HE, and *P*_5+_ was dominant for HKU1-S1. We also observed that the proportion of seronegative individuals peaked rapidly before age 1, which reflects our fitted maternal seroreversion rates and is consistent with maternal antibody kinetics for other pathogens (e.g. [23, 24]). While we did not explicitly fit age-varying seroconversion rates, these are readily recovered from serostatus-class-specific seroconversion rates and the proportion at each serostatus level by age (Figure S7). Seroconversion peaks for all sHCoVs in early childhood, reflecting the rapid loss of maternal antibodies.

**Table 1:**
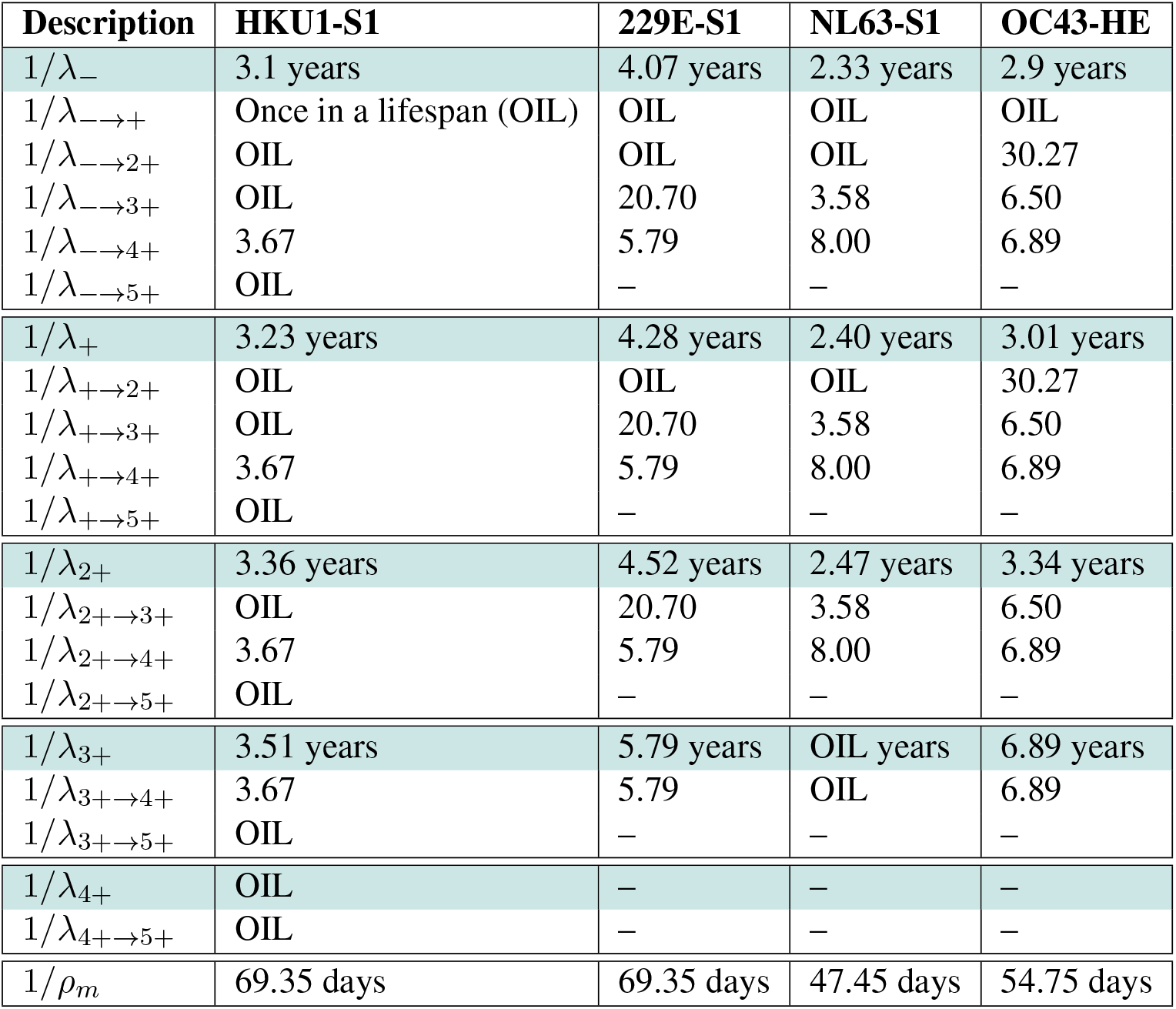
Estimates of time to seroconversion (1*/λ*) and maternal seroreversion (1*/ρ*). Values are shown for the Variation Model, in units of years or days. Blue highlighted rows show the overall time to seroconversion for a given serostatus level (e.g. 1*/λ*_−_ out of *P*_−_), calculated as the inverse of the sum of subrates out of that serostatus level. Parameters for which a once-in-a-lifetime boundary value (1*/*80 years) is estimated, are marked by “OIL”. Profiles on fit parameters are shown in Figure S6.

| Description | HKU1-S1 | 229E-S1 | NL63-S1 | OC43-HE |
| --- | --- | --- | --- | --- |
| $1/\lambda_-$ | 3.1 years | 4.07 years | 2.33 years | 2.9 years |
| $1/\lambda_{- \rightarrow +}$ | Once in a lifespan (OIL) | OIL | OIL | OIL |
| $1/\lambda_{- \rightarrow 2+}$ | OIL | OIL | OIL | 30.27 |
| $1/\lambda_{- \rightarrow 3+}$ | OIL | 20.70 | 3.58 | 6.50 |
| $1/\lambda_{- \rightarrow 4+}$ | 3.67 | 5.79 | 8.00 | 6.89 |
| $1/\lambda_{- \rightarrow 5+}$ | OIL | — | — | — |
| $1/\lambda_+$ | 3.23 years | 4.28 years | 2.40 years | 3.01 years |
| $1/\lambda_{+ \rightarrow 2+}$ | OIL | OIL | OIL | 30.27 |
| $1/\lambda_{+ \rightarrow 3+}$ | OIL | 20.70 | 3.58 | 6.50 |
| $1/\lambda_{+ \rightarrow 4+}$ | 3.67 | 5.79 | 8.00 | 6.89 |
| $1/\lambda_{+ \rightarrow 5+}$ | OIL | — | — | — |
| $1/\lambda_{2+}$ | 3.36 years | 4.52 years | 2.47 years | 3.34 years |
| $1/\lambda_{2+ \rightarrow 3+}$ | OIL | 20.70 | 3.58 | 6.50 |
| $1/\lambda_{2+ \rightarrow 4+}$ | 3.67 | 5.79 | 8.00 | 6.89 |
| $1/\lambda_{2+ \rightarrow 5+}$ | OIL | — | — | — |
| $1/\lambda_{3+}$ | 3.51 years | 5.79 years | OIL years | 6.89 years |
| $1/\lambda_{3+ \rightarrow 4+}$ | 3.67 | 5.79 | OIL | 6.89 |
| $1/\lambda_{3+ \rightarrow 5+}$ | OIL | — | — | — |
| $1/\lambda_{4+}$ | OIL | — | — | — |
| $1/\lambda_{4+ \rightarrow 5+}$ | OIL | — | — | — |
| $1/\rho_m$ | 69.35 days | 69.35 days | 47.45 days | 54.75 days |

**Figure 4:**
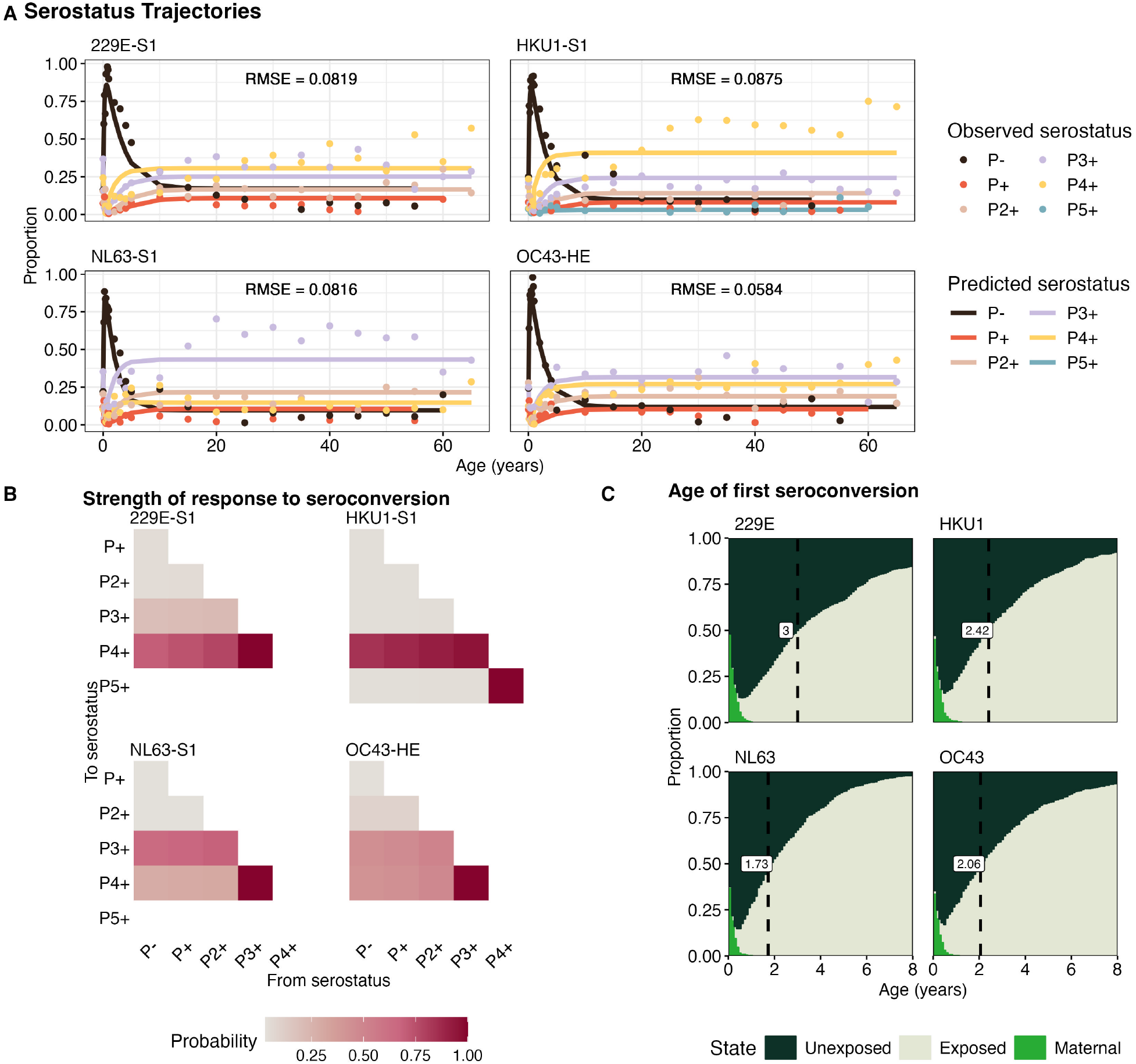
Estimates of exposure histories. (A) Predicted serostatus proportions (solid lines) vs. observed serostatus proportions (points) for the Variation Model, including RMSE values. Observed data are aggregated every 1.5 months for individuals *<* 1 year, every year for individuals *<* 5 years, and every five years for those *>*= 5 years of age. (B) Given that an individual is seroconverting out of a serostatus compartment, the probabilities of ending up in a each destination serostatus level are shown. Note that in the case of seroconverting out of the second-highest serostatus level, the probability of ending at the maximum serostatus will always be 100%. (C) Median age of first seroconversion due to infection (dashed lines) by sHCoV. These ages were inferred from the stochastic simulations with 1000 individuals for each sHCoV. The proportions of unexposed, exposed, and individuals with maternal immunity are shown. Unexosed individuals are those who are in the seronegative class *P*_−_ for the first time. Estimates of exposure histories for the Binary Model are shown in Figure S5.

We find that NL63-S1 exhibits the fastest seroconversion rate of all SHCoVs during infancy until 2.25 years of age, when OC43-HE becomes dominant throughout early childhood. At 12.75 years, NL63-S1 again has the highest seroconversion rate and this persists for the rest of the lifespan. 229E-HE showed the slowest seroconversion rate for all ages.

As serostatus levels increased, the overall time to the next seroconversion event subtly increased - meaning fewer infections are estimated to occur in higher serostatus levels. This could suggest that prior exposure is mildly protective against future infection, however these small increases in seroconversion time out of a given serostatus level were almost entirely driven by drop-off in the number of destination serostatus compartments (Table 1). We estimated the scalar change in protection between serostatus levels during fitting, and protection was unchanged for all but one sHCoV and serostatus level (Figure S6, Table 1). For example, for HKU1-S1, the overall time to seroconvert out of *P* − was 3.1 years, and the time to seroconvert out of *P*_+_ was 3.23 years, but there was no change in protection between *P* − and *P* +. That is, when looking at the sub-rates of seroconversion *into P* + through *P*_5+_, our model estimated that 1*/λ*_−⟶4+_ and 1*/λ*_+⟶4+_ were both 3.67 years, and all other sub-rates were estimated at once-in-a-lifetime (1*/*80years^−1^). The difference between 3.1 years to seroconversion vs. 3.23 years to seroconversion is, therefore, due to the drop-off of *λ*_−⟶+_. Given that increases in the time to seroconversion are driven almost completely by a decreased number of serostatus levels to convert to, this suggests the possibility that some individuals at higher serostatus levels are not boosting antibodies upon exposure. The single exception to a lack of increasing protection was observed for NL63-S1, where seroconversion from *P*_3+_ ⟶ *P*_4+_ occurred once in a lifetime, but seroconversion from *P*_2+_⟶ *P*_4+_ occurred every 8 years. However, *P*_4+_ is the maximum serostatus level attainable for NL63-S1, and therefore a low seroconversion rate into *P*_4+_ out of *P*_3+_ compared to *P*_2+_ could still be due to a lack of detectable antibody boosting rather than protection.

Next, the fine-grain seroconversion rates captured by the Variation Model allowed us to compare the probability of seroconverting from a given origin serostatus level, to a given destination serostatus level (Figure 4B). For all sHCoVs, seroconverting into *P*_+_ or *P*_2+_ was unlikely (also seen in Table 1), suggesting that these serostatus levels are more likely to be observed after an individual has seroreverted from a higher serostatus level. For 229E-S1, and OC43-HE, serostatus level *P*_4+_ was the most likely destination serostatus level when converting from any lower status, followed by *P*_3+_. For NL63-S1, *P*_3+_ was the most probable destination serostatus level, followed by *P*_4+_. For HKU1-S1, seroconversions were almost entirely concentrated into *P*_4+_.

Finally, we wanted to estimate the sHCoV exposure landscape across the lifespan. Using the Gillespie algorithm [25] with fit parameters from the serocatalytic models, we generated stochastic serostatus trajectories for each sHCoV with 1, 000 individuals. By tracking seroconversion and seroreversion times for all trajectories, we estimated numerically the number of lifetime infections, likely age of first seroconversion due to infection, as well as the dynamics of maternal antibody loss (Figures 4C, S8). The first infection after waning of maternal antibodies is likely to be NL63, occuring at a median age of 1.73 years, followed by OC43 (median 2.06 years of age), HKU1 (median 2.42 years of age) and finally 229E (median 3.00 years of age) (Figure 4C). Nearly all individuals with maternal seropositivity have lost this status by age 1, regardless of sHCoV. We estimate the average number of seroconversion events for a given sHCoV to be 10.79-13.17 throughout the lifetime (Figure S8), which is about half the number of events inferred by the Binary Model. When looking at the characteristics of individual trajectories, we observed that most individuals fluctuate between seronegative and seropositive throughout the lifespan (Figure S8). Few individuals remain lifelong seropositive after the first infection, and no individuals maintain lifelong seronegativity, consistent with our age-of-first-infection estimates (Figure S8). Thus, the maintenance of seronegative adults in Figure 4A reflects individuals who have waned after a previous exposure, rather than immunologically naive adults.

## Discussion

Serocatalytic models have traditionally been used to gain insight into pathogen dynamics from cross-sectional data [1]. Yet, these methods remain limited by the inability to capture heterogeneity in serological levels, the exposure history, or the immune response of the host, despite complex adaptations [4] and new modeling approaches [26]. Here, we used a Gaussian mixture model to characterize a gradient of serological status for the four seasonal coronaviruses and leveraged this characterization to glean new insights from serocatalytic models - using cross-sectional data. Crucially, we found that models capturing a serostatus gradient always performed better than models that only captured seronegative/seropositive dynamics, regardless of any other model assumptions tested. Moreover, the best-fit model for each virus was one that explicitly captured heterogeneity in the strength of serological response to infection.

When comparing dynamics across sHCoVs, our models suggested that HKU1 infection is likely to generate the strongest serological responses measured by optical density, and NL63 the weakest. While empirical analyses have found that pre-existing antibodies to 229E showed the least neutralizing activity compared to OC43 and NL63 [27], our model suggested that 229E has the second strongest serological responses as measured by optical density, higher than both OC43 and NL63. These differences might be explained by differences between antibody avidity vs. quantity induced by sHCoV antigens, which could be tested in the future. Diversity in antibody responses across sHCoVs might also be associated with complex evolutionary processes that are challenging to disentangle (e.g. [28, 29]). With respect to the quality of antibody responses, our results also showed that changes in protection against reinfection as serostatus increases are generally minimal for all sHCoVs. Instead, slower seroconversion out of higher serostatus levels is driven by a shrinking number of destination serostatus levels after seroconversion. Several possibilities could explain these dynamics. First, the wide distribution of OD_450_ values that we observed at high levels of serostatus suggest that individuals with pre-existing antibodies could be seroconverting without large enough changes in OD_450_ values to be detected by our models - or that there is more heterogeneity in responses to sero-conversion that what our models can explicitly capture. Second, it is possible that some individuals at high serostatus levels could get infected without a meaningful change in their antibody response, regardless of our measurement tools. Both of these possibilities point to an antibody ceiling effect, which has been previously observed for influenza virus (e.g. [30]). Our samples were collected before the COVID-19 pandemic became widespread, and target proteins exhibit minimal overlap across sHCoVs [19, 31], however other cross-reactive antibodies and T cell immunity might additionally influence patterns observed here. Despite our finding of limited protection against reinfection, exposure history might have some impact on outcomes from reinfection. For example, changes in the speed of antibody production upon reinfection have been observed for RSV and influenza virus [32, 33]; this could be tested empirically for sHCoVs in the future.

Our models suggested that the earliest sHCoV seroconversion due to exposure occurs at a median age of 1.75 years (NL63) and the latest at 3.00 years (229E). When considering the age of first infection and early-life disease dynamics, the fleeting window where most individuals have lost their maternal antibodies but have not yet been exposed to a pathogen can have consequences for childhood vaccination campaigns. While there are no seasonal coronavirus vaccines, sHCoV dynamics can inform SARS-CoV-2 vaccine timing in the future. Maternal antibodies have been known to inhibit vaccine-induced antibody responses in infants for several pathogens such as influenza virus [21]. Though cell-mediated immunity may be spared from this effect [34], antibody and T cell responses appear to play complementary roles in SARS-CoV-2 infection [35], which may lead to vaccine interference in young children. On the other hand, delaying vaccination might mean a missed opportunity to prevent infection. Currently, SARS-CoV-2 vaccination is recommended for infants 6 months and older [36]. Considering that SARS-CoV-2 is a betacoronavirus like OC43 and HKU1, and that the dynamics of early exposure estimated here are consistent across both alpha- and beta-coronaviruses, if SARS-CoV-2 continues to move toward an endemic pattern similar to that of the seasonal coronaviruses our models support this vaccine timing. Furthermore, vaccinating for SARS-CoV-2 before the age of first seasonal coronavirus infection may be prudent, as evidence for interactions between SARS-CoV-2 and other HCoV immunity remains inconsistent [27, 37–39].

While we focused on sHCoVs as a case study in this work, our approach can be broadly applied to other pathogens of interest, with potential differences in serological classification. For example, dengue differs from SARS-CoV-2 and sHCoVs in that the second infection can be associated with more severe disease [40]. Given this dynamic, we might expect that the development of serostatus levels is related to heterogeneous antibody boosting after infection, but with a higher density of serological observations at lower OD_450_ values and fewer serostatus components detected by mixture models - reflecting that a stronger antibody response might predispose individuals to severe disease and death. This sparsity can indeed be seen in DENV-2 OD_450_ values reported by other authors [41]. Here, we estimated candidate seroreversion rates from longitudinal data [22] -but longitudinal data is not essential to this process. Our methods can be applied with cross-sectional data alone for pathogens such as SARS-CoV-2, where seroreversion is better characterized (e.g. [42]), or by testing a wider range of seroreversion rates when these rates are not known. Moreover, our approach enabled us to stochastically generate individual serological trajectories, which can be validated in the future with longitudinal data collection. Crucially, our methods provide an opportunity to infer highly-detailed pathogen dynamics from cross-sectional data alone, when longitudinal data - which is definitionally resource- and time-intensive to collect - is not available.

While the findings presented here have potentially wide-ranging implications, it is important to acknowledge key limitations. First, this work leverages cross-sectional data. Increased collection of longitudinal serological data, especially for infants, will be important to clarify patterns in exposure history and serological heterogeneity reported here. Second, we did not incorporate seasonality in this work because our serocatalytic models captured population-level serology across age, which obscures seasonal signals. The seasonal pattern of sHCoVs in Hong Kong is variable by pathogen [43, 44] and the dominant sHCoV varies year to year [45]. In the future, exploring sHCoV seasonality with nonlinear transmission models might yield further insights, including for the age of first infection. Finally, we did not evaluate multi-sHCoV interactions or possible interactions with SARS-CoV-2. While these interactions might alter the outcomes of our analyses, they are beyond the scope of this work.

In conclusion, the approach presented here is highly flexible, and has the potential to be applied to other pathogens of interest. Gaussian mixture models can help to identify serological patterns beyond the seronegative/seropositive binary. The design of our multi-exposure models can be adapted to pathogens with any number of serostatus components, and the optimal model structure might differ by pathogen. Understanding the landscape of serological diversity and mechanisms underlying it can advance our understanding of pathogen ecology and best practices for public health.

## Methods

### Sample Collection

Between January and March 2020, a cross-sectional study was undertaken [19] at Guangzhou and Red Cross, Hong Kong on seroprevalence of four HCoVs in volunteers from Guangdong Women and Children Hospital and The Chinese University of Hong Kong. Plasma samples were collected from 1886 pediatric patients under 18 years old without signs of influenza-like illness as well as 528 volunteers whose age ranging from 16-67 years old. All peripheral blood samples were centrifuged at 3000 x g for 10 minutes at room temperature for plasma collection and kept at -80°C until used. All study procedures were performed after informed consent. The study was approved by the Human Research Ethics Committee at the Guangdong Women and Children Hospital (Approval number: 202101231) and The Chinese University of Hong Kong (IRB: 2020.229).

### Enzyme-linked immunosorbent assay (ELISA)

Nunc MaxiSorp ELISA plates (Thermo Fisher Scientific) were coated overnight at 4°C with 100 *µ*L of recombinant proteins at 1 *µ*g/mL in PBS. Of note, the S1 subunits of spike protein (His tag) of HCoV-229E (Seattle/USA/SC1073/2016), HCoV-HKU1 (Hong Kong/isolate N5/2006), HCoV-NL63 (Florida/UF-2/2015) and the hemagglutinin-esterase (HE) protein (His Tag) of HCoV-OC43 (Seattle/USA/SC9741/2016) were purchased from Sino Biological (China). On the next day, the plates were blocked with 100 *µ*l of Chonblock blocking/sample dilution ELISA buffer (Chondrex Inc, Redmon, US) and incubated at room temperature for 1 hour. Each human plasma sample was diluted to 1:100 in Chonblock blocking/sample dilution ELISA buffer and then added into the ELISA plates for a 2-hour incubation at 37°C. Plates were then washed thrice and incubated with horseradish peroxidase (HRP)-conjugated goat anti-human IgG antibody (Thermo Fisher Scientific) at 1:5,000 dilution for 1 hour at 37°C. After six washes with PBS containing 0.05% Tween 20, 100 *µ*L of 1-Step TMB ELISA Substrate Solution (Thermo Fisher Scientific) was added to each well. After incubation for 10 minutes, the reaction was stopped with 50 *µ*L of 2 M *H*_2_*SO*_4_ solution, and absorbance values were measured at 450 nm using a BioTek Synergy HTX Multimode Reader (Agilent).

### Classification of serostatus

We implemented Gaussian mixture models to classify serological samples across each sHCoV, using the MCLUST package in R [20]. In order to identify a seronegative/seropositive threshold value, we selected the pool of individuals younger than one year, which is expected to include both seropositive and seronegative samples due to the waning of maternal antibodies. For each sHCoV, we fixed *n* = 2 compartments and fit mixture models, estimating the seropositive/seronegative threshold as the midpoint between mean optical density (OD_450_) values in each of the two serostatus levels. In the full set of samples across age, we considered samples seropositive if they fell above the threshold value, and seronegative if they fell below it. In this way, we classified all data into seronegative or seropositive. In order to test a gradient serostatus classification, we reran mixture models on the seropositive data identified above, without specifying the number of components *n* in the model. This enabled the optimal number of seropositive compartments to be selected by the Bayesian Information Criterion (BIC), and yielded a gradient of seropositive groups for each sHCoV.

### Preparation of data and initial conditions

Seropositive samples in our data were grouped into “maternal” and “self-acquired” based on the age that the proportion of seronegative samples peaks. That is, seropositive individuals with age less than the peak are classed as “maternal” and those older than the peak are “self-acquired”. We aggregated continuous age measurements into age groups ranging in size from every 1.5 months (ages 0-1), one year (ages 1-5), or five years (age 5+), labeling by the lowest age in a group. These bin sizes were chosen to generate adequately smooth data for fitting while still capturing rapid changes in the distribution of OD_450_ measurements for young children.

The proportion of individuals in each serostatus and age group was then calculated. Fitting dynamic birth/death processes is not possible in these model formulations, because simulations progress over age since birth. Therefore, initial conditions for simulations at age 0 were based on the observed proportion of infants that were seronegative or maternally seropositive in data, and are provided in Table S6-S5.

### Estimating the seroreversion rate

To estimate the seroreversion rate for sHCoVs, we classified previously published longitudinal data for 10 individuals spanning 205.6 person-years [22] into serostatus levels using Gaussian mixture models. While we cannot determine the mapping of serostatus levels in this data with groups in the cross-sectional data due to the lack of a seronegative reference group, this allowed us to identify points in the data where an individual has likely seroconverted or seroreverted (serostatus level increased or decreased compared to their previous sample). For each individual and sHCoV, whenever a seroconversion event was followed by a seroreversion event, we took the time in years between these events. We then calculated the 5th and 95th quantiles of these values (including all sHCoVs), and the inverse values are used as upper and lower scenarios for the general seroreversion rate of all sHCoVs (Figure S3).

### Fitting and Profiling

To test and compare model performance, we fit seroconversion and maternal seroreversion rates across a range of model scenarios (Equations S1-S3, Figure 2) using maximum likelihood [46]. Models were fit to the proportion at each serostatus level in our cross sectional data, which was grouped by age, serostatus, and type (maternal vs. self-acquired serostatus). We calculated log-likelihood with the probability density function for the normal distribution.

In the Binary Model, we fit the seroconversion and maternal seroreversion parameters (Table S1). In the Ordered Model, we fit the seroconversion rate into the highest serostatus level (*λ*_*n*+_) as well as the relative changes in seroconversion rates at each serostatus level (Table S2). That is, if *λ*_*i*_ is the seroconversion rate at serostatus *i*, then *b*_*i*−1_ is the multiplicative change in seroconversion rate such that

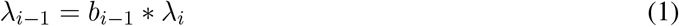

This enabled us to bound relative changes in seroconversion to be monotonically increasing with increased serostatus. In the Variation Model, we fit the set of seroconversion rates *λ*_*k*−1⟶*k*_ for all serostatuses *k*, as well as parameters {*b*_−_, *b*_+_, …, *b*_*n*+_} that modulate the change in seroconversion at each serostatus level (Table S3). In particular, we used the following scaling:

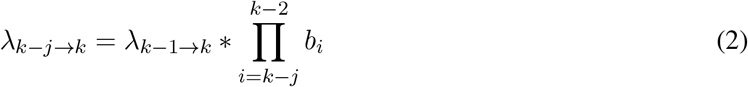

For all models, we additionally fit the standard deviation *σ* used in calculating log-likelihood.

During fitting, we assumed that each possible seroconversion event must occur at a rate at least 1/80 years^−1^ (approximately once in a lifetime [47]), and no more frequently than once per month. In order to further assure parameter identifiability, we assumed that seroconversion rates must either stay the same or decrease with increasing serostatus, informed by the fact that antibody titers for SARS-CoV-2 correlate with increased protection [48]. After fitting each scenario to the data, we selected the best-performing scenario for each sHCoV using the Akaike Information Criterion (AIC), and profiled the parameters.

For each sHCoV we tested a set of 10 distinct scenarios across three axes:

1. Models: Binary, Ordered, Variation
2. Waning of serostatus: sequential (*P*_*i*_ ⟶ *P*_*i*−1_) or direct (*P*_*i*_ ⟶ *P*_−_)
3. Seroreversion rate: 5th vs. 95th quantile

### Stochastic Trajectories

For each sHCoV and best-fit model (Binary and Variation), we generated *n* = 1000 stochastic individual trajectories using the rates of seroreversion and seroconversion found during fitting. Each individual simulation was initialized by probabilistically selecting a seronegative or maternally seropositive state according to Tables S6-S5. Then, the next event and the time until this event occurs were selected using the Gillespie algorithm [25]. We classified these trajectories into lifelong seronegative (excluding maternal seropositivity), lifelong seropositive, and fluctuating. Additionally, we estimated the total number of lifetime seroconversion events and age of first seroconversion post-waning of maternal antibodies.

## Data Availability

Upon publication, all code and data needed to evaluate this study's conclusions and reproduce figures will be made available in a permanent repository.

## Acknowledgements

SLL, ANMK, NCW, and PPM were funded by the Dr. Gert Ehrlich Cross-field Innovation Fund from Carle Illinois College of Medicine, University of Illinois Urbana-Champaign (UIUC). SLL was also funded by the Isabel Norton Award from the School of Integrative Biology, UIUC. JY was funded by the UIUC Professor Alice Carlene Helm Scholarship. The authors would like to thank the Biocluster at the Carl R. Woese Institute for Genomic Biology at UIUC for access to computing resources. We would also like to thank Dr. James O’Dwyer and Kenneth Jops, as well as the O’Dwyer Lab at UIUC for invaluable feedback on this work.

## Author Contributions

SLL, JY, HL, ST, ANMK, NCW, and PPM conceptualized the study. SLL, ANMK, NCW, and PPM acquired funding. SLL, JY, HL, ST, ANMK, NCW, and PPM contributed to methodology. Investigation: SLL, JY, HL, YWH, QT, TP, RL, ABG, EKS, LT. Software: SLL and JY. Formal analysis: SLL, JY, HL, PPM. Data curation: SLL, JY, HL, CKPM. Visualization: SLL, JY, PPM. Supervision: SLL, NCW, PPM. Project administration: SLL. Writing - original draft: SLL, JY, HL, PPM. All authors contributed to manuscript review and editing.

## Competing Interests

The authors report no competing interests.

## Notes

### Competing Interest Statement

The authors have declared no competing interest.

### Author Declarations

The study was approved by the Human Research Ethics Committee at the Guangdong Women and Children Hospital (Approval number: 202101231) and The Chinese University of Hong Kong (IRB: 2020.229)

## References

[1] James A Hay, Isobel Routledge, and Saki Takahashi. Serodynamics: a review of methods for epidemiological inference using serological data. OSF Preprints, (kqdsn), 2023.

[2] Leonhard Held, Niel Hens, Philip D O’Neill, and Jacco Wallinga. Handbook of infectious disease data analysis. CRC Press, 2019.

[3] Bryan T Grenfell and RM Anderson. The estimation of age-related rates of infection from case notifications and serological data. Epidemiology & Infection, 95(2):419–436, 1985.

[4] Neil M Ferguson, Christl A Donnelly, and Roy M Anderson. Transmission dynamics and epidemiology of dengue: insights from age–stratified sero–prevalence surveys. Philosophical Transactions of the Royal Society of London. Series B: Biological Sciences, 354(1384):757–768, 1999.

[5] A. De Thoisy, T. Woudenberg, S. Pelleau, F. Donnadieu, L. Garcia, L. Pinaud, L. Tondeur, A. Meola, L. Arowas, N. Clement, M. Backovic, M.-N. Ungeheuer, A. Fontanet, M. White, T. Woudenberg, S. Pelleau, L. Pinaud, L. Tondeur, M.-N. Ungeheuer, and A. D. Koffi. Seroepidemiology of the seasonal human coronaviruses nl63, 229e, oc43 and hku1 in france. Open Forum Infectious Diseases, 10(7), 2023.

[6] E. M. Rees, N. R. Waterlow, R. Lowe, and A. J. Kucharski. Estimating the duration of seropositivity of human seasonal coronaviruses using seroprevalence studies. Wellcome Open Research, 6:138, 2021.

[7] Eleanor M Rees, Colleen L Lau, Mike Kama, Simon Reid, Rachel Lowe, and Adam J Kucharski. Estimating the duration of antibody positivity and likely time of leptospira infection using data from a cross-sectional serological study in fiji. PLoS neglected tropical diseases, 16(6):e0010506, 2022.

[8] Bruce G Link and Jo Phelan. Social conditions as fundamental causes of disease. Journal of health and social behavior, pages 80–94, 1995.

[9] James O Lloyd-Smith, Sebastian J Schreiber, P Ekkehard Kopp, and Wayne M Getz. Superspreading and the effect of individual variation on disease emergence. Nature, 438(7066):355–359, 2005.

[10] Ruian Ke, Pamela P Martinez, Rebecca L Smith, Laura L Gibson, Agha Mirza, Madison Conte, Nicholas Gallagher, Chun Huai Luo, Junko Jarrett, Ruifeng Zhou, et al. Daily longitudinal sampling of sars-cov-2 infection reveals substantial heterogeneity in infectiousness. Nature microbiology, 7(5):640–652, 2022.

[11] Rachel J Oidtman, Philip Arevalo, Qifang Bi, Lauren McGough, Christopher Joel Russo, Diana Vera Cruz, Marcos Costa Vieira, and Katelyn M Gostic. Influenza immune escape under heterogeneous host immune histories. Trends in microbiology, 29(12):1072–1082, 2021.

[12] M. Desforges, A. Le Coupanec, P. Dubeau, A. Bourgouin, L. Lajoie, M. Dubé, and P. J. Talbot. Human coronaviruses and other respiratory viruses: Underestimated opportunistic pathogens of the central nervous system? Viruses, 12(1):14, 2019.

[13] Melisa M Shah, Amber Winn, Rebecca M Dahl, Krista L Kniss, Benjamin J Silk, and Marie E Killerby. Seasonality of common human coronaviruses, united states, 2014–2021. Emerging Infectious Diseases, 28(10):1970, 2022.

[14] Sangshin Park, Yeonjin Lee, Ian C Michelow, and Young June Choe. Global seasonality of human coronaviruses: a systematic review. In Open forum infectious diseases, volume 7, page ofaa443. Oxford University Press US, 2020.

[15] Jeffrey Shaman and Marta Galanti. Will sars-cov-2 become endemic? Science, 370(6516):527–529, 2020.

[16] J. J. Guthmiller and P. C. Wilson. Remembering seasonal coronaviruses. Science, 370(6522):1272–1273, 2020.

[17] Stephen M Kissler, Christine Tedijanto, Edward Goldstein, Yonatan H Grad, and Marc Lipsitch. Projecting the transmission dynamics of sars-cov-2 through the postpandemic period. Science, 368(6493):860–868, 2020.

[18] Jeffrey P Townsend, Hayley B Hassler, April D Lamb, Pratha Sah, Aia Alvarez Nishio, Cameron Nguyen, Alexandra D Tew, Alison P Galvani, and Alex Dornburg. Seasonality of endemic covid-19. MBio, 14(6):e01426–23, 2023.

[19] Y. Luo, H. Lv, S. Zhao, Y. Sun, C. Liu, C. Chen, W. Liang, K. Kwok, Q. W. Teo, R. T. So, Y. Lin, Y. Deng, B. Li, Z. Dai, J. Zhu, D. Zhang, J. Fernando, N. C. Wu, H. M. Tun, and X. Mu. Age-related seroprevalence trajectories of seasonal coronaviruses in children including neonates in guangzhou, china. International Journal of Infectious Diseases, 127:26–32, 2023.

[20] Luca Scrucca, Chris Fraley, T. Brendan Murphy, and Adrian E. Raftery. Model-Based Clustering, Classification, and Density Estimation Using mclust in R. Chapman and Hall/CRC, 2023.

[21] Stefan Niewiesk. Maternal antibodies: clinical significance, mechanism of interference with immune responses, and possible vaccination strategies. Frontiers in immunology, 5:446, 2014.

[22] Arthur WD Edridge, Joanna Kaczorowska, Alexis CR Hoste, Margreet Bakker, Michelle Klein, Katherine Loens, Maarten F Jebbink, Amy Matser, Cormac M Kinsella, Paloma Rueda, et al. Seasonal coronavirus protective immunity is short-lasting. Nature medicine, 26(11):1691–1693, 2020.

[23] Atila Kılıc, Sevin Altınkaynak, Vildan Ertekin, and Tacettin Inandı. The duration of maternal measles antibodies in children. Journal of tropical pediatrics, 49(5):302–305, 2003.

[24] Elke Leuridan, Niel Hens, Veronik Hutse, Marc Aerts, and Pierre Van Damme. Kinetics of maternal antibodies against rubella and varicella in infants. Vaccine, 29(11):2222–2226, 2011.

[25] Daniel T Gillespie. Exact stochastic simulation of coupled chemical reactions. The journal of physical chemistry, 81(25):2340–2361, 1977.

[26] V. Yman, M. T. White, J. Rono, B. Arcà, F. H. Osier, M. Troye-Blomberg, S. Boström, R. Ronca, I. Rooth, and A. Färnert. Antibody acquisition models: A new tool for serological surveillance of malaria transmission intensity. Scientific Reports, 6(1), 2016.

[27] Jan Lawrenz, Qinya Xie, Fabian Zech, Tatjana Weil, Alina Seidel, Daniela Krnavek, Lia van der Hoek, Jan Münch, Janis A Müller, and Frank Kirchhoff. Severe acute respiratory syndrome coronavirus 2 vaccination boosts neutralizing activity against seasonal human coronaviruses. Clinical Infectious Diseases, 75(1):e653–e661, 2022.

[28] James R Otieno, Joshua L Cherry, David J Spiro, Martha I Nelson, and Nídia S Trovão. Origins and evolution of seasonal human coronaviruses. Viruses, 14(7):1551, 2022.

[29] Wendy K Jo, Christian Drosten, and Jan Felix Drexler. The evolutionary dynamics of endemic human coronaviruses. Virus Evolution, 7(1):veab020, 2021.

[30] Robert M Jacobson, Diane E Grill, Ann L Oberg, Pritish K Tosh, Inna G Ovsyannikova, and Gregory A Poland. Profiles of influenza a/h1n1 vaccine response using hemagglutination-inhibition titers. Human vaccines & immunotherapeutics, 11(4):961–969, 2015.

[31] Andrew R Crowley, Harini Natarajan, Andrew P Hederman, Carly A Bobak, Joshua A Weiner, Wendy Wieland-Alter, Jiwon Lee, Evan M Bloch, Aaron AR Tobian, Andrew D Redd, et al. Boosting of cross-reactive antibodies to endemic coronaviruses by sars-cov-2 infection but not vaccination with stabilized spike. Elife, 11:e75228, 2022.

[32] Robert C Welliver, Tej N Kaul, Theodore I Putnam, Martha Sun, Katherine Riddlesberger, and Pearay L Ogra. The antibody response to primary and secondary infection with respiratory syncytial virus: kinetics of class-specific responses. The Journal of pediatrics, 96(5):808–813, 1980.

[33] James A Hay, Karen Laurie, Michael White, and Steven Riley. Characterising antibody kinetics from multiple influenza infection and vaccination events in ferrets. PLoS Computational Biology, 15(8):e1007294, 2019.

[34] Marjolein RP Orije, Kirsten Maertens, Véronique Corbière, Nasamon Wanlapakorn, Pierre Van Damme, Elke Leuridan, and Françoise Mascart. The effect of maternal antibodies on the cellular immune response after infant vaccination: a review. Vaccine, 38(1):20–28, 2020.

[35] Jingyi Yan, Chandrashekar Ravenna Bangalore, Negin Nikouyan, Sofia Appelberg, Daniela Nacimento Silva, Haidong Yao, Anna Pasetto, Friedemann Weber, Sofie Weber, Olivia Larsson, et al. Distinct roles of vaccine-induced sars-cov-2-specific neutralizing antibodies and t cells in protection and disease. Molecular Therapy, 32(2):540–555, 2024.

[36] Centers for Disease Control and Prevention. Covid-19 vaccination for people who are pregnant or breastfeeding.

[37] M. Dugas, T. Grote-Westrick, U. Merle, M. Fontenay, A. E. Kremer, F. Hanses, R. Vollenberg, E. Lorentzen, S. Tiwari-Heckler, J. Duchemin, S. Ellouze, M. Vetter, J. Fürst, P. Schuster, T. Brix, C. M. Denkinger, C. Müller-Tidow, H. Schmidt, P.-R. Tepasse, and J. Kühn. Lack of antibodies against seasonal coronavirus oc43 nucleocapsid protein identifies patients at risk of critical covid-19. Journal of Clinical Virology, 139:104847, 2021.

[38] P. R. Wratil, N. A. Schmacke, B. Karakoc, A. Dulovic, D. Junker, M. Becker, U. Rothbauer, A. Osterman, P. M. Spaeth, A. Ruhle, M. Gapp, S. Schneider, M. Muenchhoff, J. C. Hellmuth, C. Scherer, J. Mayerle, M. Reincke, J. Behr, S. Käb, and O. T. Keppler. Evidence for increased sars-cov-2 susceptibility and covid-19 severity related to pre-existing immunity to seasonal coronaviruses. Cell Reports, 37(13):110169, 2021.

[39] Elizabeth M Anderson, Eileen C Goodwin, Anurag Verma, Claudia P Arevalo, Marcus J Bolton, Madison E Weirick, Sigrid Gouma, Christopher M McAllister, Shannon R Christensen, JoEllen Weaver, et al. Seasonal human coronavirus antibodies are boosted upon sars-cov-2 infection but not associated with protection. Cell, 184(7):1858–1864, 2021.

[40] Hsin-I Shih, Yu-Ching Wang, Yu-Ping Wang, Chia-Yu Chi, and Yu-Wen Chien. Risk of severe dengue during secondary infection: A population-based cohort study in taiwan. Journal of Microbiology, Immunology and Infection, 57(5):730–738, 2024.

[41] Barbara Batista Salgado, Fábio Carmona de Jesus Maués, Maele Jordão, Renato Lemos Pereira, Daniel A Toledo-Teixeira, Pierina L Parise, Fabiana Granja, Higo Fernando Santos Souza, Marcio Massao Yamamoto, Jannifer Oliveira Chiang, et al. Antibody cross-reactivity and evidence of susceptibility to emerging flaviviruses in the dengue-endemic brazilian amazon. International Journal of Infectious Diseases, 129:142–151, 2023.

[42] Michael Loesche, Elizabeth W Karlson, Opeyemi Talabi, Guohai Zhou, Natalie Boutin, Rachel Atchley, Gideon Loevinsohn, Jun Bai Park Chang, Mohammad A Hasdianda, Adetoun Okenla, et al. Longitudinal sars-cov-2 nucleocapsid antibody kinetics, seroreversion, and implications for seroepidemiologic studies. Emerging infectious diseases, 28(9):1859, 2022.

[43] Susanna KP Lau, Patrick CY Woo, Cyril CY Yip, Herman Tse, Hoi-wah Tsoi, Vincent CC Cheng, Paul Lee, Bone SF Tang, Chris HY Cheung, Rodney A Lee, et al. Coronavirus hku1 and other coronavirus infections in hong kong. Journal of clinical microbiology, 44(6):2063–2071, 2006.

[44] Susan S Chiu, Kwok Hung Chan, Ka Wing Chu, See Wai Kwan, Yi Guan, Leo Lit Man Poon, and JSM Peiris. Human coronavirus nl63 infection and other coronavirus infections in children hospitalized with acute respiratory disease in hong kong, china. Clinical infectious diseases, 40(12):1721–1729, 2005.

[45] Wai-Sing Chan, Siu-Kei Yau, Man-Yan To, Sau-Man Leung, Kan-Pui Wong, Ka-Chun Lai, Ching-Yan Wong, Chin-Pang Leung, Chun-Hang Au, Thomas Shek-Kong Wan, et al. The seasonality of respiratory viruses in a hong kong hospital, 2014–2023. Viruses, 15(9):1820, 2023.

[46] Ben Bolker and R Development Core Team. bbmle: Tools for General Maximum Likelihood Estimation, 2023. R package version 1.0.25.1.

[47] World Bank Group. Life expectancy at birth, total (years) - china, hong kong sar, china. https://data.worldbank.org/indicator/SP.DYN.LE00.IN?locations=CN-HK.

[48] David S Khoury, Timothy E Schlub, Deborah Cromer, Megan Steain, Youyi Fong, Peter B Gilbert, Kanta Subbarao, James A Triccas, Stephen J Kent, and Miles P Davenport. Correlates of protection, thresholds of protection, and immunobridging among persons with sars-cov-2 infection. Emerging Infectious Diseases, 29(2):381, 2023.

[49] NH Barton. The ecological detective: Confronting models with data. by ray hilborn and marc mangel. princeton university press, 1997. 315+ xvii pages. price£ 30.00 cloth,£ 16.95 paper. isbn 0 691 03496 6; 0 691 03497 4 (pbk). Genetics Research, 70(2):175–181, 1997.

