## Supplementary Materials for "Reimagining the serocatalytic model for infectious diseases: a case study of common coronaviruses"

### S1 Supplementary text

#### Serocatalytic Models

##### Traditional Serocatalytic Model

In order to capture binary serostatus dynamics, we first added maternal seropositivity to a traditional serocatalytic model [1] which we call the "Binary Model" (BM) (eq. S1). Individuals start as maternally seropositive $M_1$ and wane at a rate $\rho_m$ to seronegative $P_-$, or start directly in the seronegative group. Individuals move from the $P_-$ class to $P_+$ via seroconversion due to exposure at rate $\lambda$, and serorevert to $P_-$ due to antibody waning at rate $\rho$.

$$\begin{aligned}
\frac{dP_-}{da} &= -\lambda P_- + \rho_m M_+ + \rho P_+ \\
\frac{dP_+}{da} &= \lambda P_- - \rho P_+ \\
\frac{dM_+}{da} &= -\rho_m M_+
\end{aligned} \tag{S1}$$

##### Adapted Serocatalytic Models

We next adapted the Binary Model to include multiple seropositive compartments for each sHCoV, corresponding to the optimal serostatus gradient identified by our Gaussian mixture models. In this model, which we denote the "Ordered Model" (eq. S2), individuals start in $P_-$ (seronegative) or maternally seropositive ($M_i$), with eventual seroreversion $\rho_m$ from $M_i$ directly to $P_-$. Once maternal seropositivity has waned, individuals can seroconvert sequentially into higher serostatus - $P_i \to P_{i+1}$ - at a rate $\lambda_i$. Seroreversion is also sequential, with serostatus waning from $P_i$ to $P_{i-1}$ at a rate $\rho_{i,i-1}$ that is constant across sHCoVs and serostatus levels.

$$\begin{aligned}
\frac{dP_-}{da} &= \rho_{+,-}P_+ - \lambda_- P_- + \rho_m \left( \sum M_{i+} \right) \\
\frac{dP_+}{da} &= \rho_{2+,+}P_{2+} + \lambda_- P_- - \lambda_+ P_+ - \rho_{+,-}P_+ \\
\frac{dM_+}{da} &= -\rho_m M_+ \\
&\vdots \\
\frac{dP_{n+}}{da} &= \lambda_{(n-1)+}P_{(n-1)+} - \rho_{n+,(n-1)+}P_{n+} \\
\frac{dM_{n+}}{da} &= -\rho_m M_{n+}
\end{aligned} \tag{S2}$$

While the Ordered Model is more flexible than the Binary Model, it does not account for heterogeneity in an individual's response to a single exposure. To capture this possibility, we tested a serocatalytic model where individuals could seroconvert from $P_i$ to any higher serostatus level $P_{i+k}$, to account that individuals might respond differently to a single exposure. We call this the "Variation Mode" (eq. S3).

$$
\begin{aligned}
\frac{dP_-}{da} &= \rho_{+,-}P_+ - (\lambda_{-\to+} + \lambda_{-\to 2+} + \lambda_{-\to 3+} + \cdots + \lambda_{-\to n+})P_- + \rho_m\left(\sum M_{i+}\right) \\
\frac{dP_+}{da} &= \rho_{2+,+}P_{2+} + \lambda_{-\to+}P_- - (\lambda_{+\to 2+} + \cdots + \lambda_{+\to n+})P_+ - \rho_{+,-}P_+ \\
\frac{dM_+}{da} &= -\rho_m M_+ \\
&\vdots \\
\frac{dP_{n+}}{da} &= (\lambda_{(n-1)+\to n+}P_{(n-1)+} + \cdots + \lambda_{-\to n+}P_-) - \rho_{n+,(n-1)+}P_{n+} \\
\frac{dM_{n+}}{da} &= -\rho_m M_{n+}
\end{aligned}
\tag{S3}
$$

For each of the Ordered and Variation models, we also implemented versions where seroreversion moves individuals directly from $P_i$ to $P_-$ at a rate $\rho_{i,-}$, rather than seroreverting sequentially.

#### Profiling algorithm

To profile parameters for these best-fit scenarios, we implemented an algorithm that takes successively smaller steps away from the fit parameter value, aiming to approximate the threshold $T$ = loglik $- 1.92$ and thus determine the 95% confidence interval on the parameter [49]. To find the upper boundary of a parameter, we proceed as follows:

1. If the parameter is already on the user-specified boundary of permissible parameter values, stop. The upper boundary is indeterminate. Otherwise, proceed to step 2.
2. Fix the profiling parameter at test parameter = parameter estimate + $s$, where $s$ is the fit parameter value or $0.001$, whichever is greater. Rerun the maximum likelihood estimate to fit all other parameters.
   (a) If the new log likelihood value is *smaller* than the threshold $T$ (we have gone too far) or adding the step size to the parameter would exceed the user-specified parameter boundary, divide the step size by 10 and retest: test parameter = parameter estimate + $s/10$.
   (b) If the new log likelihood value is still *greater* than the threshold $T$ (we have not gone far enough), add the step size again (e.g. test parameter = parameter estimate + $2s$ and retest.
   (c) If the new log likelihood value is exactly the threshold $T$, stop and return the tested parameter value.
3. Continue iterating with additional step sizes/smaller step sizes until the threshold is reached or dividing by 10 causes the step size to be $\leq 10^{-5}$. At this stage, there are two possibilities:
   (a) The exact parameter value resulting in likelihood $T$ has been determined.

(b) Adding $10^{-4}$ to the fit parameter value resulted in a likelihood smaller than the threshold $T$, triggering division by 10 and the end of the profiling. Then the threshold likelihood is achieved by a parameter value $p$ such that parameter estimate $< p <$ parameter estimate $+ 10^{-4}$.

Having either identified the true upper boundary of the confidence interval or estimated to within $10^{-4}$ of the true value, we then profiled the lower boundary by the same method, subtracting the step size instead of adding it.

### S2 Supplementary Tables

| **Parameter** | **Meaning** |
|---|---|
| $\lambda_-$ | Seroconversion |
| $\rho_m$ | Maternal seroreversion |
| $\sigma$ | Standard deviation |

Table S1: Fit parameters in the Binary Model. Parameter start values and boundary values are listed in Table S4.

| **Parameter** | **Meaning** |
|---|---|
| $\lambda_{(n-1)+}$ | Seroconversion into $n+$ serostatus |
| $b_{n-2}$ | Ratio of $\frac{\lambda_{(n-2)+}}{\lambda_{(n-1)+}}$ |
| $\vdots$ | $\vdots$ |
| $b_0$ | Ratio of $\frac{\lambda_-}{\lambda_+}$ |
| $\rho_m$ | Maternal seroreversion |
| $\sigma$ | Standard deviation |

Table S2: Fit parameters in the Ordered Model. Here, $n$ represents the total number of seropositive components.Parameter start values and boundary values are listed in Table S4.

| **Parameter** | **Meaning** |
|---|---|
| $\lambda_{(n-1)+\to n+}$ | Seroconversion from $(n-1)+$ to $n+$ serostatus |
| $\lambda_{(n-2)+\to(n-1)+}$ | Seroconversion from $(n-2)+$ to $(n-1)+$ |
| $\vdots$ | $\vdots$ |
| $\lambda_{-\to+}$ | Seroconversion from $-$ to $+$ |
| $b_{i-1}$ | Ratio of $\frac{\lambda_{(i-1)\to j}}{\lambda_{i\to j}}$, for $j \in (1, n)$, $1 < i < j$ |
| $\rho_m$ | Maternal seroreversion |
| $\sigma$ | Standard deviation |

Table S3: Fit parameters in the Variation Model. Here, $n$ represents the total number of seropositive components. Each $\lambda_k$ is calculated as the sum of $\lambda_{i\to k}$, $i < k$. Parameter start values and boundary values are listed in Table S4.

| Model | Parameter | Start | Lower | Upper |
|---|---|---|---|---|
| Binary | $\sigma$ | 0.2 | 0.001 | 5 |
| | $\rho_m$ | 1 | 0 | 365 |
| | $\lambda_-$ | 1 | 1/80 | 12 |
| Ordered | $\sigma$ | 0.2 | 0.001 | 5 |
| | $\rho_m$ | 1 | 0 | 365 |
| | $b_0$ | 1 | 1 | 8000 |
| | $b_1$ | 1 | 1 | 8000 |
| | $b_2$ | 1 | 1 | 8000 |
| | $\lambda_3$ | 1 | 1/80 | 12 |
| (HKU1-S1 only) | $b_3$ | 1 | 1 | 8000 |
| (HKU1-S1 only) | $\lambda_4$ | 1 | 1/80 | 12 |
| Variation | $\sigma$ | 0.2 | 0.001 | 5 |
| | $\rho_m$ | 1 | 0 | 365 |
| | $b_0$ | 1 | 1 | 8000 |
| | $b_1$ | 1 | 1 | 8000 |
| | $b_2$ | 1 | 1 | 8000 |
| | $\lambda_{-\to+}$ | 1 | 1/80 | 12 |
| | $\lambda_{+\to2+}$ | 1 | 1/80 | 12 |
| | $\lambda_{2+\to3+}$ | 1 | 1/80 | 12 |
| | $\lambda_{3+\to4+}$ | 1 | 1/80 | 12 |
| (HKU1-S1 only) | $b_3$ | 1 | 1 | 8000 |
| (HKU1-S1 only) | $\lambda_{4+\to5+}$ | 1 | 1/80 | 12 |

Table S4: Starting values and user-specified boundaries for maximum likelihood estimation.

| sHCoV | P- | M+ |
|---|---|---|
| HKU1-S1 | 25.00 | 75 |
| NL63-S1 | 22.06 | 77.94 |
| OC43-HE | 24.26 | 75.74 |
| 229E-S1 | 16.18 | 83.82 |

Table S5: Initial conditions for binary classification models: proportion in each serostatus level.

| sHCoV | P- | M+ | M2+ | M3+ | M4+ | M5+ |
|---|---|---|---|---|---|---|
| HKU1-S1 | 25.00 | 8.09 | 18.38 | 21.32 | 23.53 | 3.68 |
| NL63-S1 | 22.06 | 11.76 | 20.59 | 35.29 | 10.29 | – |
| OC43-HE | 24.26 | 10.29 | 27.94 | 25.00 | 12.50 | – |
| 229E-S1 | 16.18 | 7.35 | 15.44 | 36.76 | 24.26 | – |

Table S6: Initial conditions for gradient classification models: proportion in each serostatus level.

| **Parameter** | **sHCOV** |
| --- | --- |
| $b_0$ | All |
| $b_1$ | All |
| $b_2$ | 229E, HKU1, OC43 |
| $b_3$ | HKU1 |
| $\lambda_{-\rightarrow+}$ | All |
| $\lambda_{+\rightarrow 2+}$ | 229E, HKU1, NL63 |
| $\lambda_{2+\rightarrow 3+}$ | HKU1 |
| $\lambda_{3+\rightarrow 4+}$ | NL63 |
| $\lambda_{4+\rightarrow 5+}$ | HKU1 |

Table S7: Parameters for which the lower confidence interval could not be identified due to user specified boundaries (Table S4)

### S3 Supplementary Figures

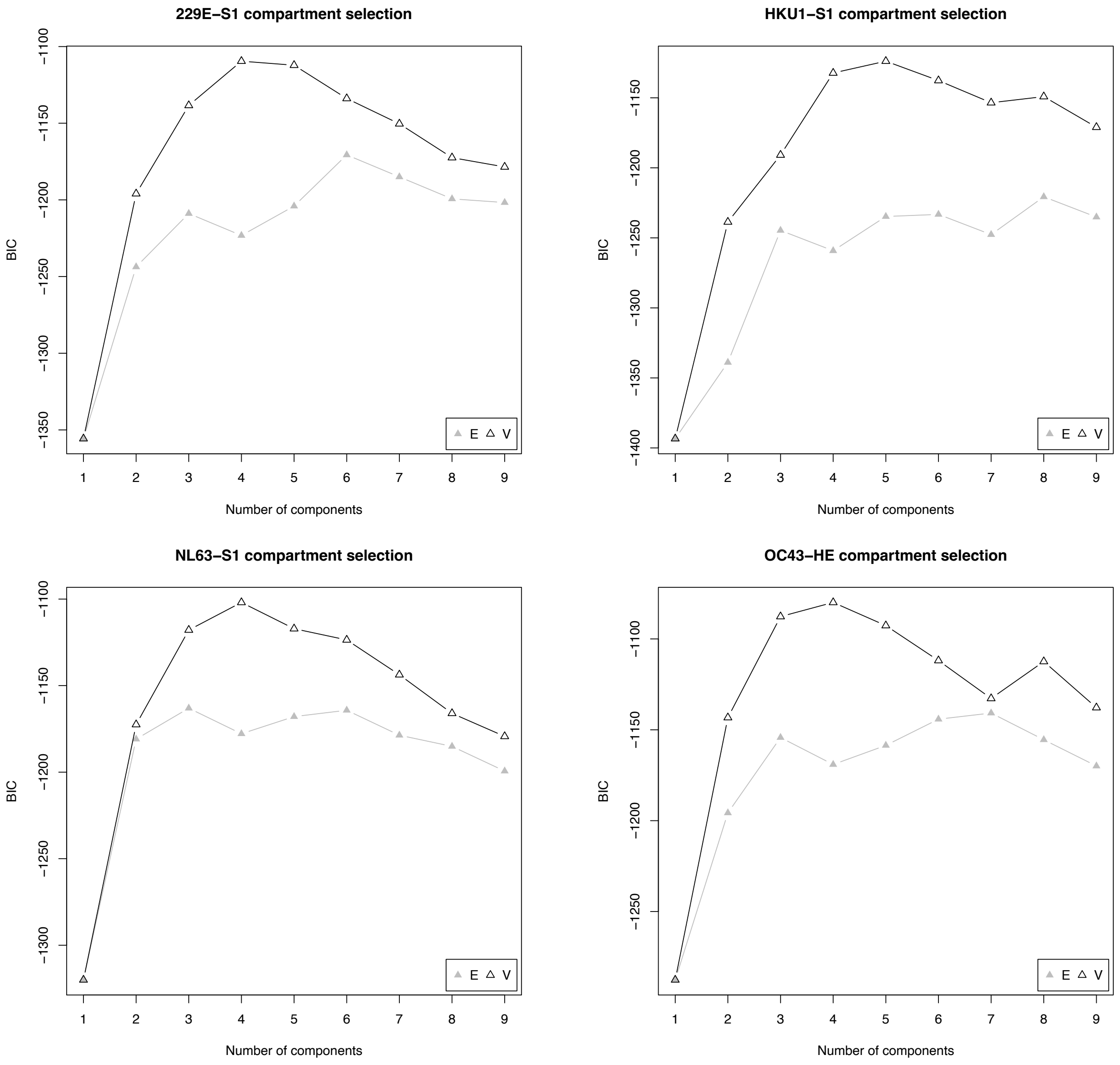


Figure S1: Model selection. Model selection is performed using mclust [20] for each sHCoV, across number of seropositive components. “E” and “V” are univariate models with (E)qual variance and (V)ariable variance respectively. In mclust, BIC is calculated using the following formula: $2L - n \cdot \log(n)$ where $n$ is the number of components and $L$ is the log likelihood.

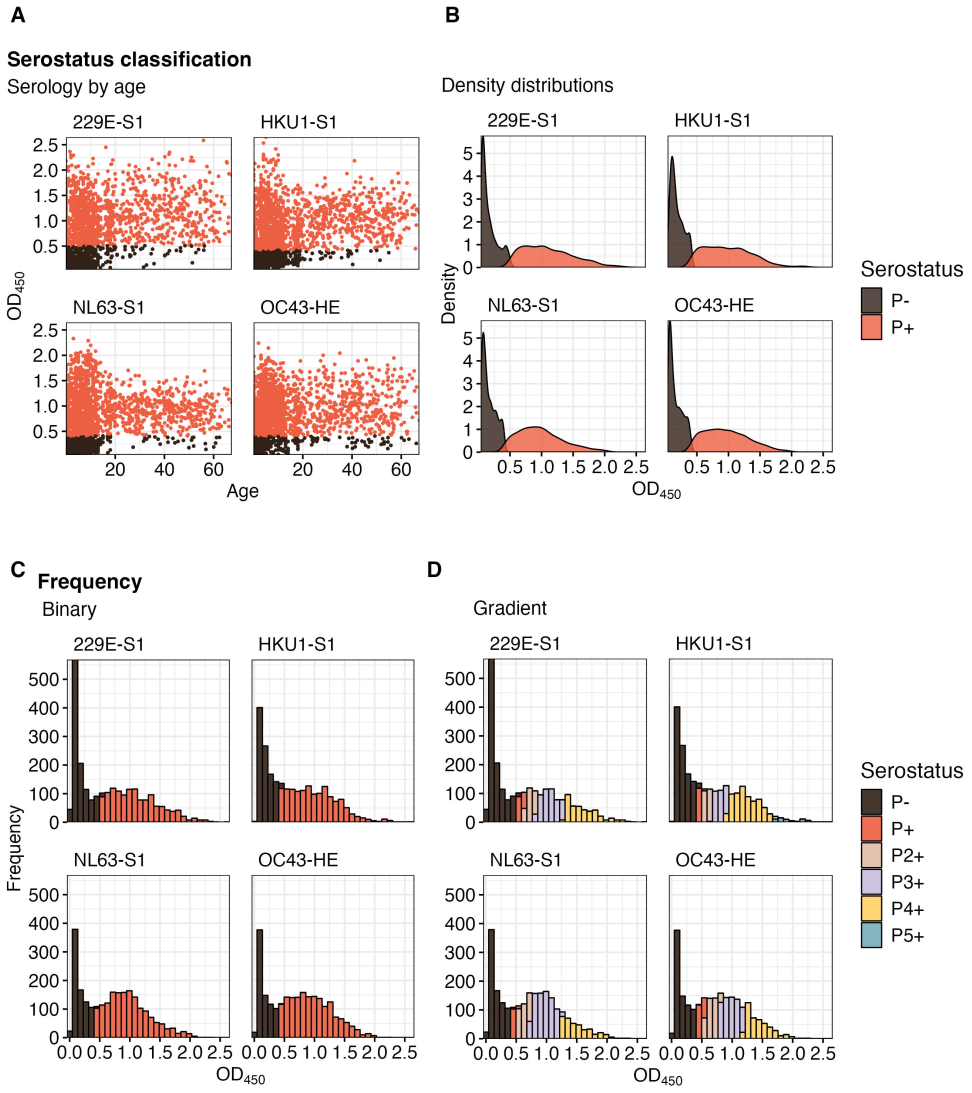


Figure S2: Serostatus classification. (A) $OD_{450}$ values by age under binary classification, colored by serostatus level. (B) $OD_{450}$ probability density under binary classification ($n = 2,414$). The color of density curves indicates the serostatus level as classified by the Gaussian mixture model. (C-D) Frequency of $OD_{450}$ values, shown under binary (C) or gradient (D) classifications and colored by serostatus level. In (C-D), $OD_{450}$ values are aggregated into 30 bins.

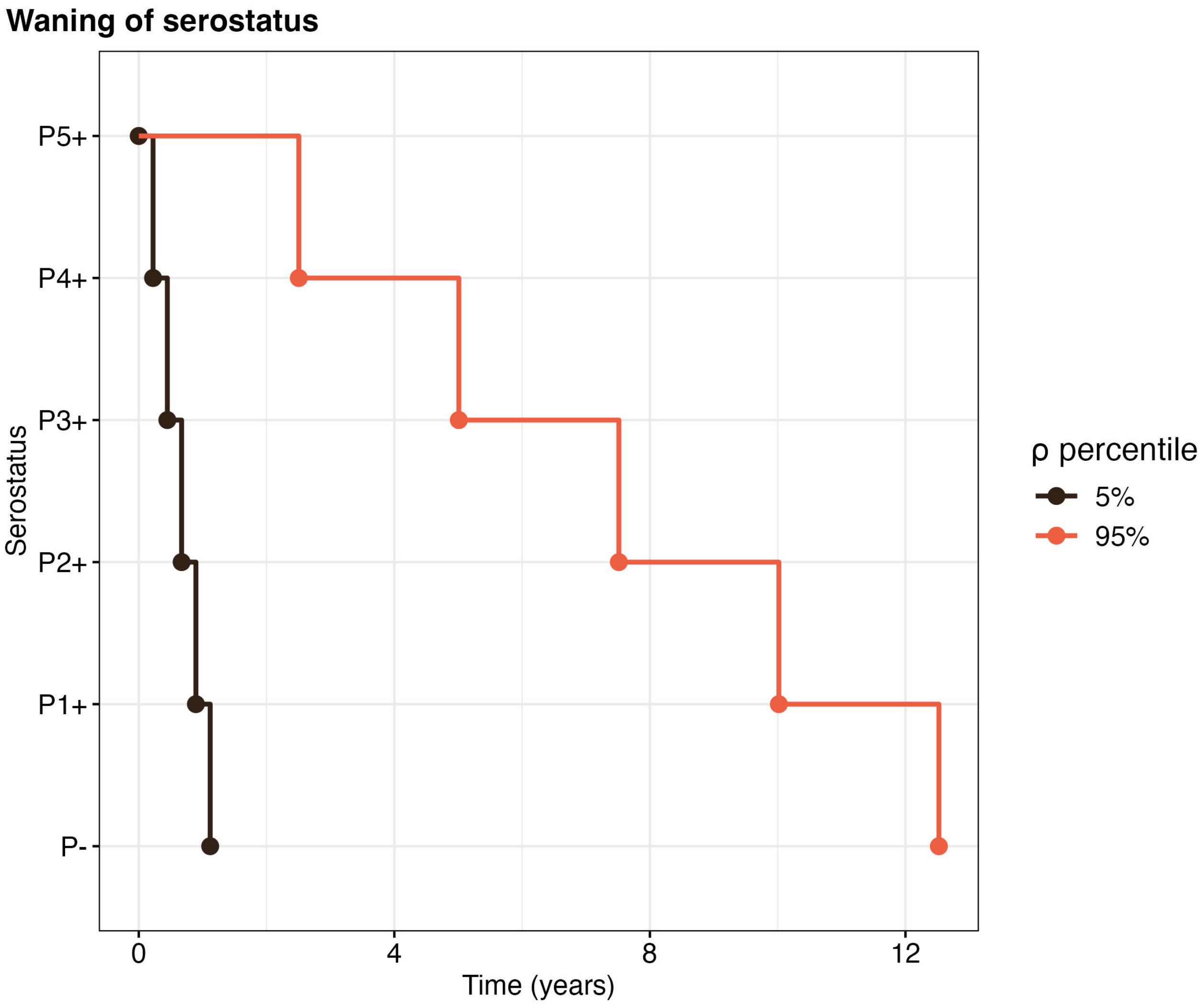


Figure S3: Waning of serostatus levels. Seroreversion trajectories assuming no new seroconversions, for the 5th (black) and 95th (red) percentiles of seroreversion rate estimated from [22].

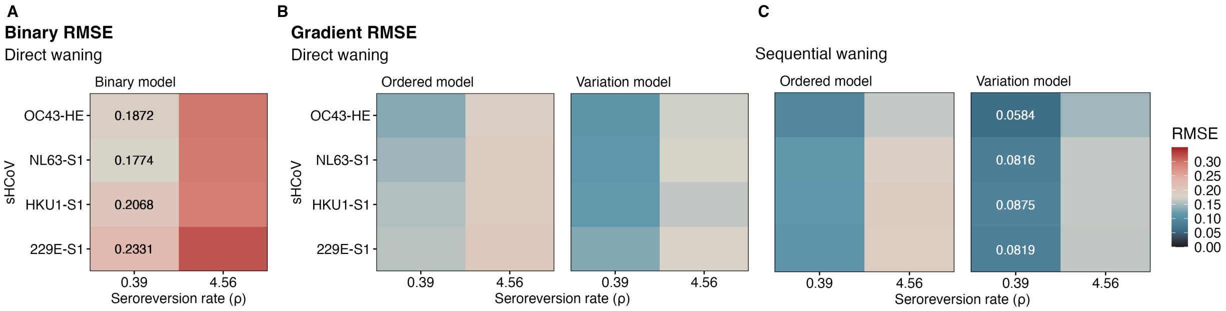


Figure S4: Goodness of fit. (A) RMSE values for the Binary Model. Two values for the seroreversion rate $\rho$ are tested [22]. Lower RMSE values indicate a better model fit, and the best (white text) and worst (black text) RMSE values are shown for each sHCoV. Fit parameters are shown in Tables S1, starting parameters and boundary values are shown in Table S4, and initial conditions are shown in Table S5. (B-C) RMSE values across the Ordered and Gradient models, with direct (B) or sequential (C) waning. Two values for the seroreversion rate $\rho$ are tested [22]. Lower RMSE values indicate a better model fit, and the RMSE values of the best fit model are shown for each sHCoV (white text). Fit parameters are shown in Tables S2-S3, starting parameters and boundary values are shown in Table S4, and initial conditions are shown in Table S6.

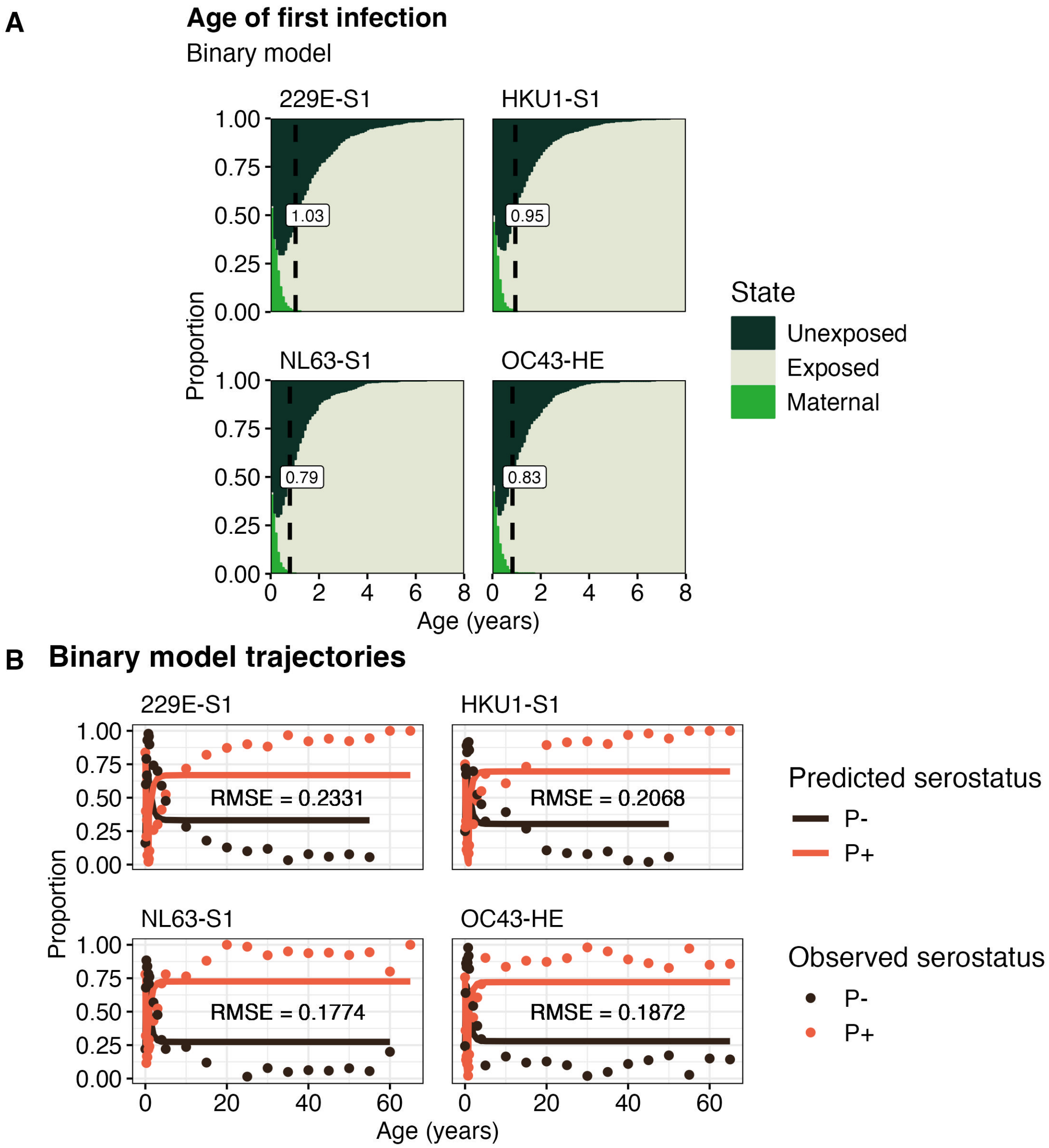


Figure S5: Estimates of exposure histories. (A) Predicted serostatus (solid lines) vs. observed serostatus (points) for the best fit Binary Models are shown, including RMSE values. Observed data are aggregated every 1.5 months for individuals $<$ 1 year, every year for individuals $<$ 5 years, and every five years for those $>=$ 5 years of age. (B) Median age of first infection (dashed lines) by sHCoV in the Binary Model ($n$ = 1000 stochastic individual simulations). The proportion unexposed, exposed, or with maternal immunity is shown.

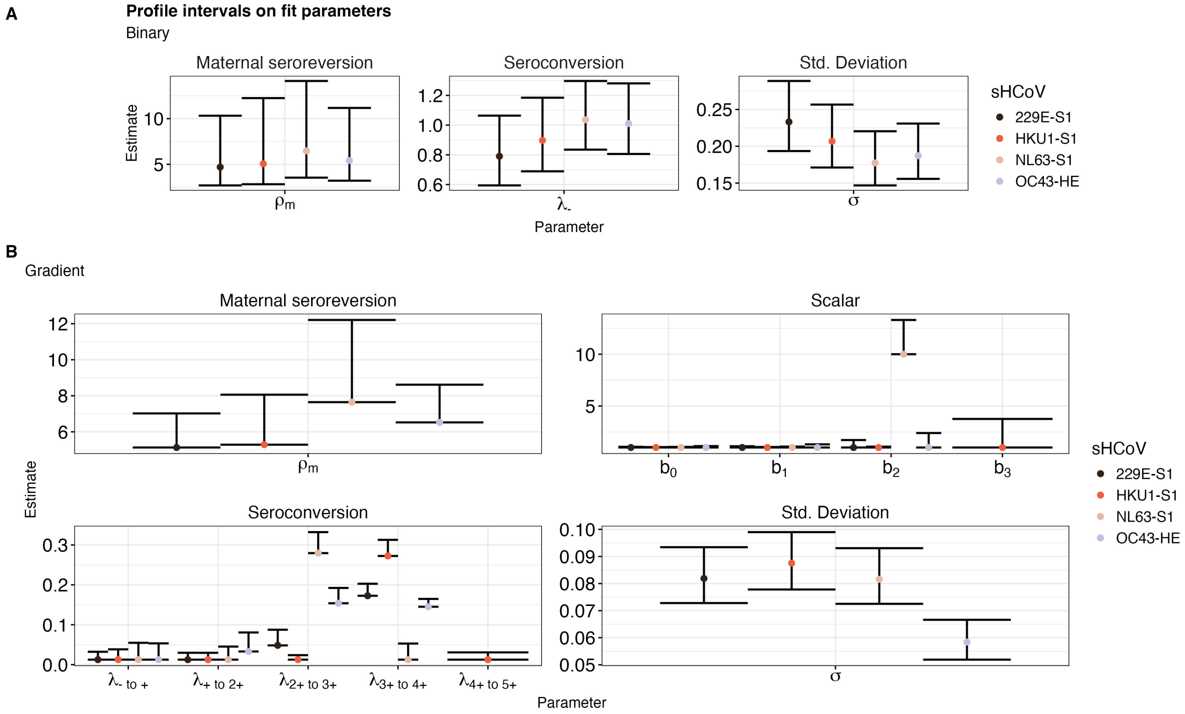


Figure S6: Parameter profiles. (A) 95% confidence intervals (whiskers) for fit parameter values (dots) in the best-fit Binary Model ($\rho = 0.39$), calculated by profiling. All upper and lower values were identified. (B) 95% confidence intervals and boundary values (whiskers) for fit parameters (dots) in the best-fit Variation Model ($\rho = 0.39$), calculated by profiling. All upper confidence values were identified. Where profiling lower bounds was not possible due to prespecified bounds for fitting (Table S4), we reported the lower parameter bound. These parameters are listed in Table S7.

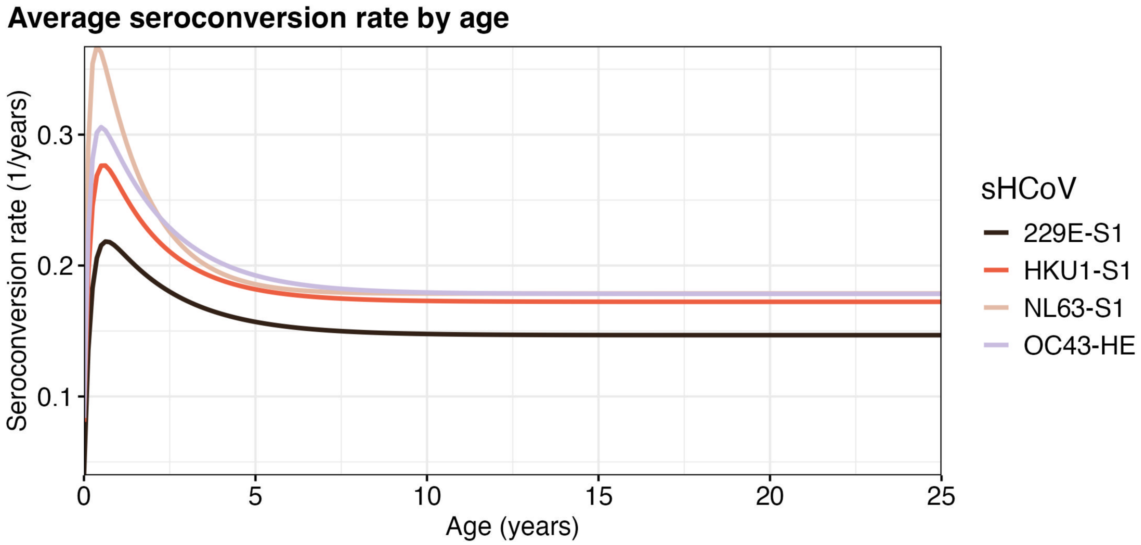


Figure S7: Seroconversion by age. Age-varying seroconversion rates, calculated as a weighted average of seroconversion rates for each serostatus level. Rates are weighted by the proportion in each group at each age (Figure 4A).

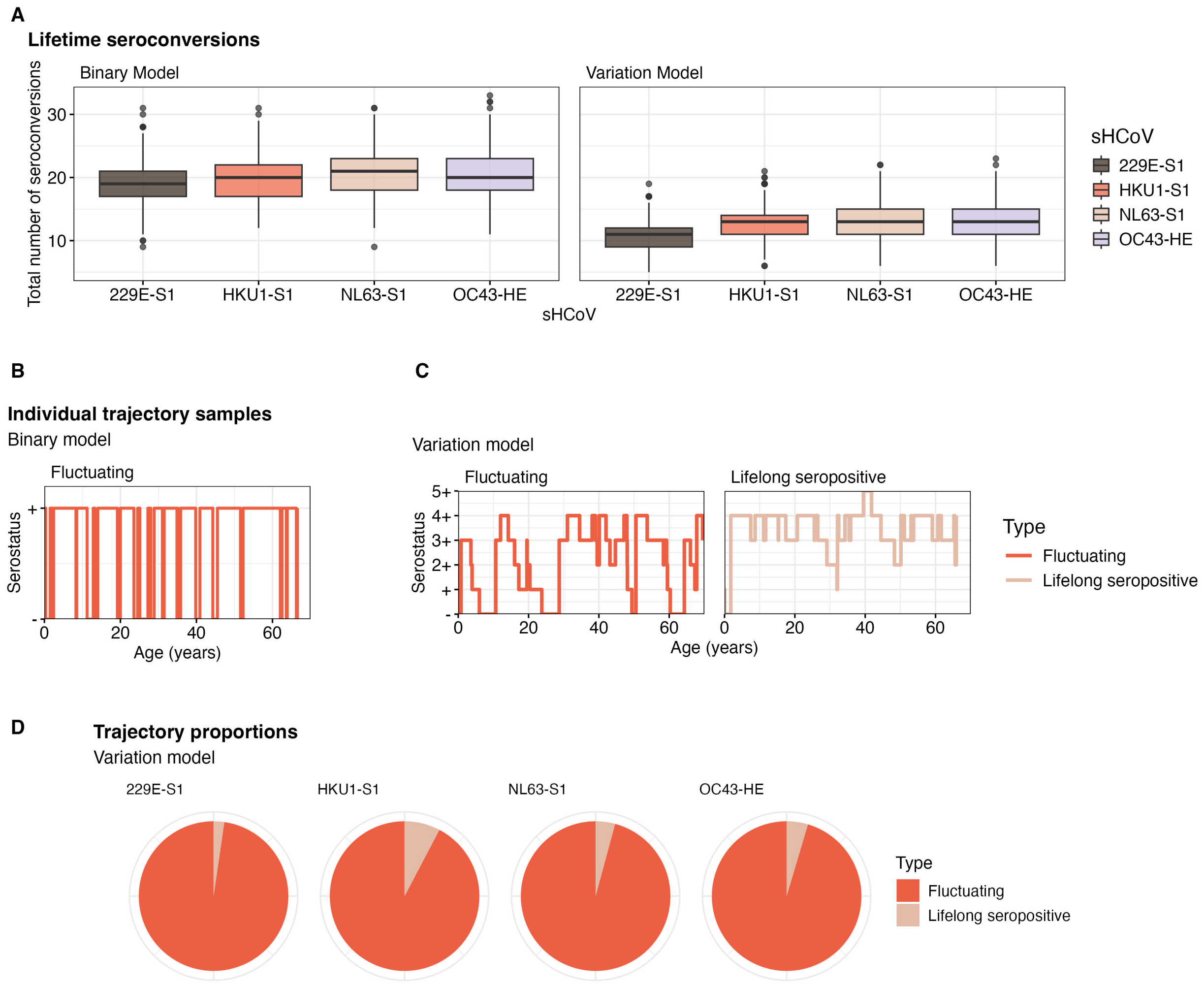


Figure S8: Stochastic trajectories. (A) Number of lifetime seroconversions in the Binary and Variation models, from $n = 1000$ individual trajectories per sHCoV. (B-C) Sample individual trajectories in the best fit Binary (B) and Variation (C) models. "Fluctuating" trajectories are those that return to seronegative at least once after first infection. "Lifelong Seropositive" are those that stay seropositive throughout the lifespan, after the first infection. In (B), trajectory 17 is shown for OC43-HE. In (C), trajectory 98 of OC43-HE is shown for "Fluctuating", and trajectory 821 for HKU1-S1 is shown for "Lifelong Seropositive". (D) Proportion of trajectories in the Variation Model that are "Fluctuating" or "Lifelong Seropositive" ($n = 1000$ simulations per sHCoV).
